# Compliance to 4Ds of antimicrobial stewardship practices in a tertiary care centre

**DOI:** 10.1101/2021.03.17.21253814

**Authors:** Diksha Dixit, Rajat Ranka, Prasan Kumar Panda

## Abstract

**Background:** Indiscriminate use of antimicrobials has been a cause of concern worldwide in recent years because of the potential to give rise to antimicrobial resistance and difficult to treat infections. Antimicrobial Stewardship describes the practice of promoting the selection of the right drug, dosage, delivery route, and duration of antimicrobial therapy (4Ds) to curtail these concerns. Nevertheless, it is important to quantify the magnitude of the problem in terms of the percentage of adherence with respect to each of the 4Ds described above. This will aid in the identification of areas for potential interventions to improve use.

**Methods:** We undertook a prospective review of the medical records of patients admitted in the medicine wards of a tertiary care centre in Northern India. All those patients who were prescribed on antimicrobials were included and their records reviewed for the indication, drug, dose, delivery, and duration (or by asking the treating physician if not documented). Adherence to the guidelines was determined by referring to the updated literature (local, national, or international standard treatment guidelines or textbooks) for each specific disease condition.

**Results:** Of the total 304 patients, the choice of drugs was appropriate and matched the guidelines in 218 (72%) patients. Adherence to the right dose in 210 (69%), route of delivery in 216 (71%), and to duration in 197 (65%) were observed. Full adherence to all the above mentioned parameters was observed in 196 (64.5%). Maximum adherence was observed in the treatment of skin and soft tissue infections (100 %) while minimum adherence was observed while administering medical prophylaxis (40 %). Among the non-adherents, over-prescription was observed in 10 (3.3 %), under-prescription in 12 (3.9 %), choice was inappropriate in 46 (15.1 %) and in 30 (10 %) patients, antimicrobials were not indicated but prescribed.

**Conclusion:** Internist practices antimicrobial stewardship with respect to the prevalent guidelines for right prescription in 2/3^rd^ admitted patients, a sub-optimal one. Right drug, dose, delivery route, duration of therapy are practiced in 72%, 69%, 71%, and 65% patients respectively. To increase adherence to 100%, bedside stewardship practices in form of prospective audit and feedback must be improved.

## Background

Discovery of antimicrobials has revolutionized the practice of medicine, making previously so called ‘lethal’ infections readily treatable. Like all medications, antimicrobials have serious side effects including (paradoxically) many difficult to treat conditions like *Clostridium difficile* infection (1). Misuse of antimicrobials has contributed to the antimicrobial resistance, which has become one of the most serious and growing threat to the public health globally. It is implicated from the fact that recently a superbug expressing NDM1 gene has been found in the arctic region (2). Potential for spread of resistant organisms means that their unjustified use can also affect the health of those who are not even exposed to them.

A growing body of evidence demonstrates that hospital based programs dedicated to optimizing antimicrobial use, commonly referred to as “antimicrobial stewardship program (ASP),” can both optimize the treatment of infections and reduce adverse events associated with antimicrobial use. This led CDC to enforce all hospitals in US to have ASP in 2014 (3). However till now in India, it is not even being practiced in few institutions. Among many interventions of ASP, prospective audit and feedback regarding adherence to standard treatment guidelines becomes important in guiding physicians to prescribe all appropriate antimicrobials, to avoid unjustified prescription, to reduce the emergence of resistant microbes, to support high quality clinical practice, and to minimize unnecessary expenses (4).The implementation of evidence-based guidelines for use of antimicrobials has been shown to improve the overall patient outcome. However, many previous studies from outside India have shown that adherence to policy recommendations has been suboptimal averaging 40% (5).

Even in India, prescription pattern monitoring studies (PPMS) conducted at various places have concluded inappropriate use of antimicrobials and lack of adherence to standard treatment guidelines (6). Therefore, first step in improving rational use of antimicrobials is to understand prescribing patterns. This will aid in the identification of areas for potential interventions to improve use.

Prescription patterns are assessed to determine the adherence to right antimicrobial drug against the stated condition (diagnosis), dose, delivery route, duration and de-escalation as soon as possible. This is a popular concept in ASP – commonly known as the 5Ds of optimal antimicrobial therapy (7). The aim of the present study is to measure the adherence (or compliance) of treating physicians’ practicing patterns of these Ds based on the antimicrobial guidelines in a tertiary care hospital.

To report our findings, we followed the STROBE (Strengthening the Reporting of Observational Studies in Epidemiology) guidelines (8).

## Materials and methods

### Study setting and design

The study was a prospective review of medical records of admitted patients in the Department of Internal Medicine at a tertiary care hospital in North India. It was conducted over a six month period, from September 2019 to February 2020.

### Study population

Review of records was done for only those in-patients who were prescribed on antimicrobials. We undertook the prevalence of antimicrobial prescribing in the Medicine ward to be 51% based on a 2014 Indian study (9). Thus, it required a sample size of 304 with 95% confidence and a precision of 0.2.

### Methodology

After institutional ethical committee approval of the study, data on antimicrobials was collected from patient’s medical records and reviewed against the indication, drug name, dose, delivery route, and duration of antimicrobial (or by asking the treating physician if not properly documented). The treating physician was also asked regarding the guidelines he/she has referred to, for each patient and documented in the proforma. Flexibility was kept regarding the choice of guidelines (as long as they were deemed to be from a reputed authority) and any particular single guideline was not chosen as reference as it was assumed that different clinicians may opt to refer to different guidelines for their patients. The study flow followed is shown (Fig. 1)

**Figure 1:**
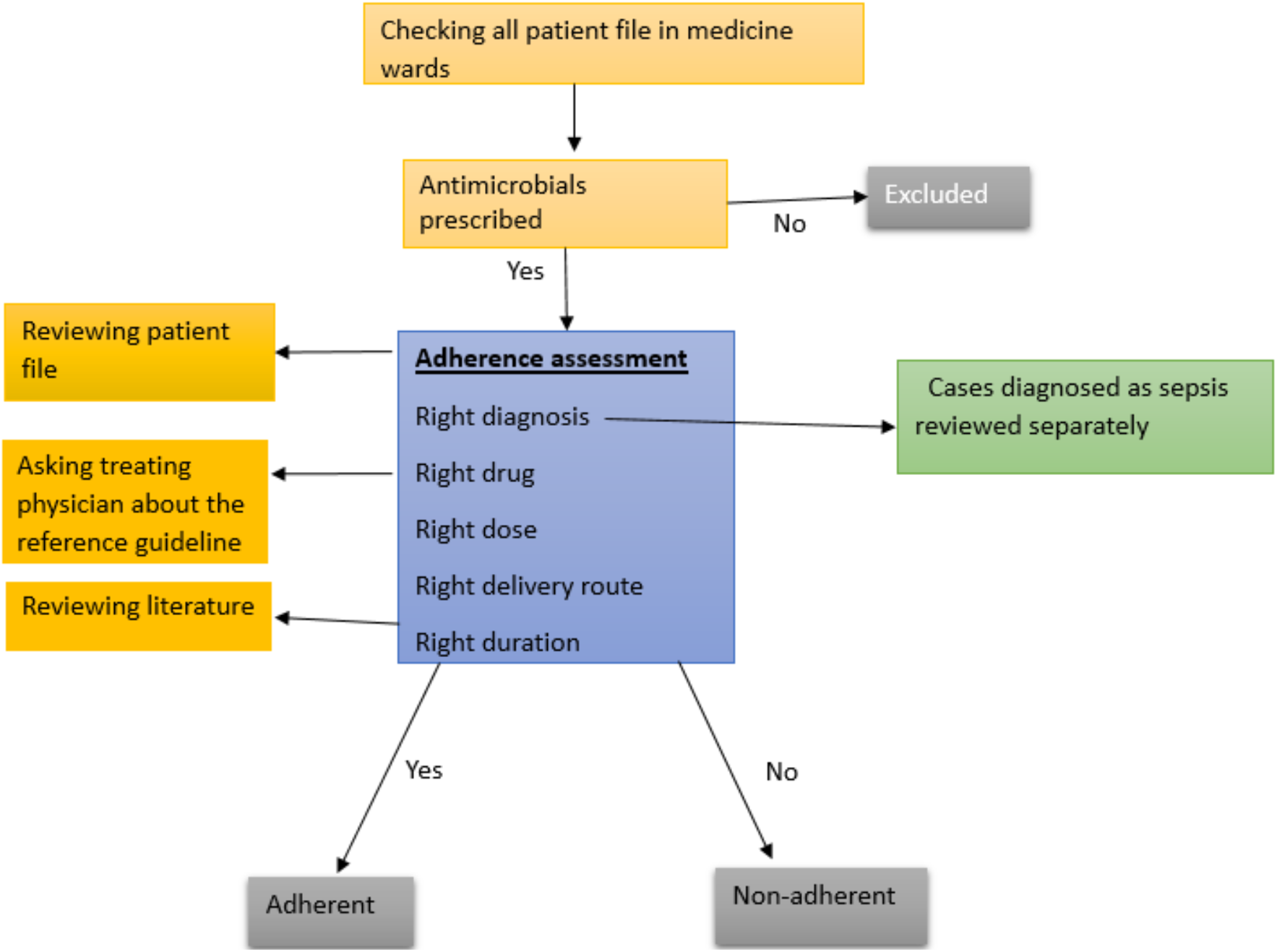
The study flow of antimicrobial prescription adherence

The main outcome of the study was the level of adherence/compliance to antimicrobial guidelines. Adherence to the guidelines was determined by referring to the updated literature (local, national, or international standard treatment guidelines and textbooks) for each specific disease condition. For each patient, detailed discussion among investigators was done regarding the diagnosis approach and management based on literature review and then adherence was entered in the proforma. The diagnosis method used by treating physicians were assumed right (100% adherence) for two reasons: firstly, if diagnosis is not right, then no point of adherence checking w.r.t. other Ds; secondly, the diagnostic approach is different from physician to physician. And right duration and de-escelation of the antimicrobial were combined assessed as fourth D since both are complementary to each other. The pattern of non-adherence was also identified and documented under four categories: over-prescription, under-prescription, choice not correct, and antimicrobial not indicated. At any time, decision of the investigators was not communicated to practicing physicians (i.e. no feedback) to get real practice scenarios.

### Data Analysis

After obtaining the required data in Microsoft Excel® sheet, they were evaluated for completeness, analysed, and interpreted. Frequency and proportions were calculated.

## Results

### Basic characteristics

A total of 304 hospitalized prescriptions were assessed. All the prescriptions were initiated by medical officers (junior residents/senior residents/Faculty). Out of 304 prescriptions, 155 were of females and 149 were of males. Median age was 46.88 years. For better understanding, indications of antimicrobial prescriptions were recorded as infection syndromes rather than individual disease (Fig. 2). Most physicians quoted the following guidelines - Indian Council of Medical Research (ICMR), National Institute for Health and Care Excellence (NICE), Infectious Diseases Society of America (IDSA), AIIMS Antimicrobial Policy, and Harrison’s Textbook of Internal Medicine. For the classification of drugs we used the ‘Guidelines for ATC classification and DDD assignment’ by WHO (10). Beta-lactams were the most frequently prescribed antimicrobial category and were included in 197 prescriptions (64.8 %). Amongst them, penicillins were prescribed in 96 (31.6 %) and other beta-lactam antimicrobials (cephalosporins, carbapenems, and monobactams) were prescribed in 101 (33.2 %) cases. Macrolides and lincosamides were included in 59 (19.4 %) prescriptions. Quinolones as well as first line ATTs were both included in 23 (7.5 %) prescriptions each, followed by antifungals in 22 (7.2 %) and tetracyclines in 20 prescriptions (6.6 %). Antivirals were prescribed for 7 (2.3 %) patients and aminoglycosides in only 3 (1 %) patients.

**Figure 2:**
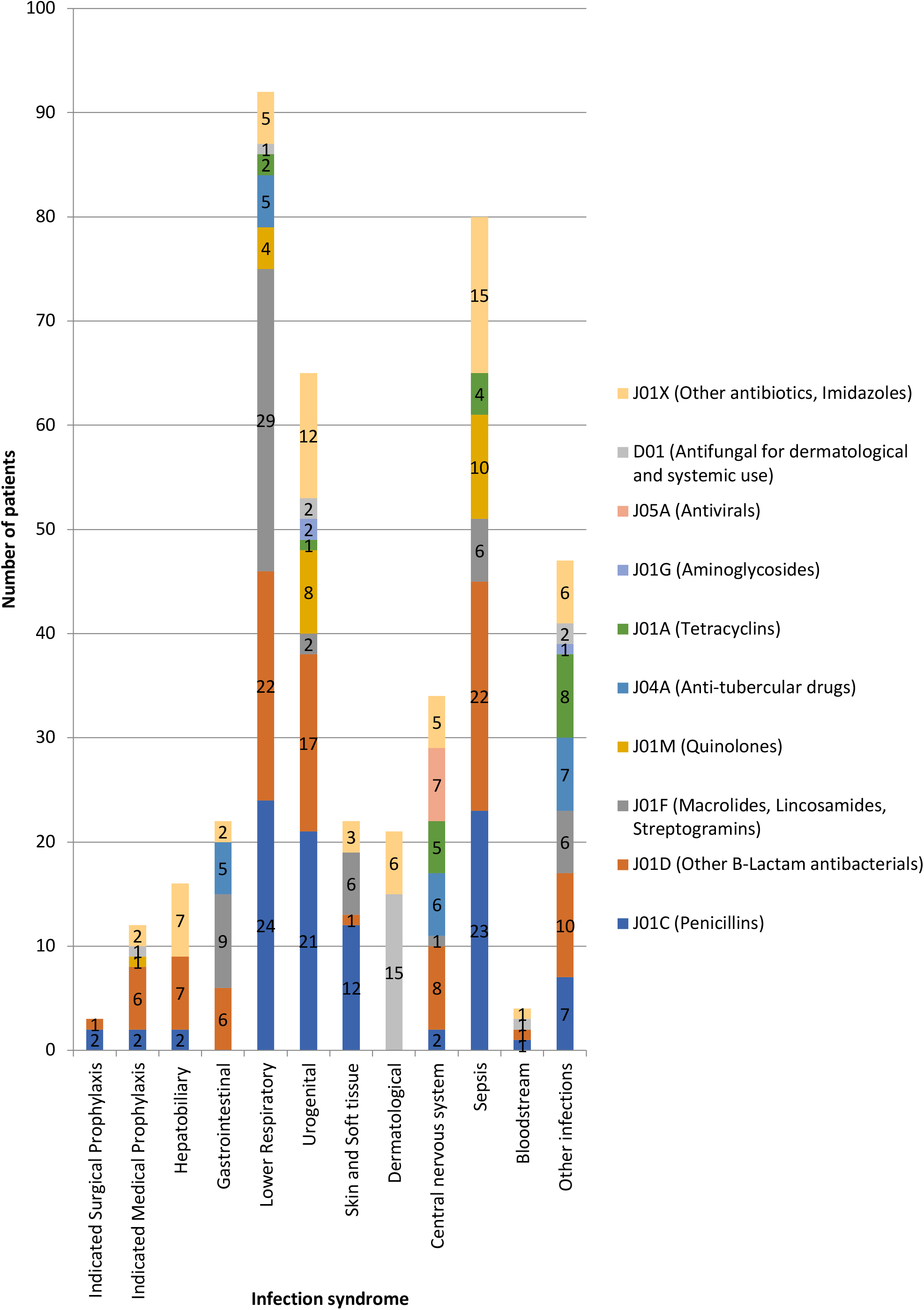
Distribution of antimicrobial prescriptions by indication.

### Compliance to guidelines in antimicrobial prescribing

Fully adherent (compliant) antimicrobial prescriptions were in 196 (64.5%) patients. In 3 (1%) prescriptions, adherence couldn’t be assessed due to lack of standard treatment guidelines for the diagnosed indication. Out of 304 prescriptions, choice of antimicrobials was appropriate and matched the guidelines in 218 (72%). Out of those 218, adherence to the route of delivery was observed in 216 (71%), to dose in 210 (69%) and to duration in 197 (65%) prescriptions (Table 1). There were 7 cases where the diagnosis/indication for starting of antimicrobials was unclear even after interviewing the treating physician and hence they have been included in the non-adherent category. In 10% prescriptions, antimicrobials were not indicated but prescribed, while in 15% prescriptions, there was incorrect choice. Over-prescription and under-prescription were in 3.3 % and 3.9 % respectively (Table 2). Respiratory tract infections was the leading indication for antimicrobial prescribing followed by urinary tract infections (both comprised of 1/3^rd^ prescriptions).

**Table 1:** Level of adherence to the guidelines as per category of indication for antimicrobial prescription.

|  |  |  |  |  |  |  |
| --- | --- | --- | --- | --- | --- | --- |
| Category of indication# | Total indicated cases | Proper adherence to |  |  |  | % Full adherence in each category ( number in parenthesis) |
|  |  | Right drug | Right dose | Right delivery route | Right duration |  |
| Indicated Surgical Prophylaxis | 3 | 3 | 2 | 2 | 2 | 66.6% (2) |
| Indicated Medical Prophylaxis | 15 | 8 | 6 | 8 | 6 | 40% (6) |
| Bloodstream infections | 2 | 2 | 2 | 2 | 2 | 100% (2) |
| **Infections involving** |  |  |  |  |  |  |
| Hepatobiliary System | 10 | 8 | 8 | 8 | 6 | 60% (6) |
| Gastrointestinal system | 20 | 17 | 17 | 17 | 17 | 85% (17) |
| Respiratory tract infections | 67 | 39 | 36 | 38 | 36 | 53.7% (36) |
| Urinary tract infections | 50 | 37 | 37 | 37 | 29 | 58% (29) |
| Central Nervous system | 29 | 26 | 26 | 26 | 23 | 79.3% (23) |
| Skin & Soft tissue infections | 16 | 16 | 16 | 16 | 16 | 100% (16) |
| Other Dermatological indications | 14 | 12 | 12 | 12 | 11 | 78.5% (11) |
| Others† | 24 | 19 | 18 | 19 | 18 | 75% (18) |
| Sepsis syndrome with defined focus | 47 | 31 | 30 | 31 | 31 | 63.8 % (30) |
| **Total\*** | **304** | **218** | **210** | **216** | **197** | **64.5 % (196)** |
| **\* Includes antimicrobials for unclear diagnosis/conditions:** 7 (where it could not be ascertained for what indication, the antibiotic has been started)<br>**† Includes infection of sinuses, rickettsia infections (scrub typhus), rheumatic heart disease, conditions involving febrile neutropenia, acute febrile illnesses, disseminated tuberculosis and histoplasmosis.**<br>**# Details of category of indication for antimicrobial prescription includes:**<br>○ Indicated surgical prophylaxis: prophylaxis for snake bite and excisional biopsy<br>○ Indicated medical prophylaxis: prophylaxis for spontaneous bacterial peritonitis, acute rheumatic fever, active GI bleed, and hospital acquired infections in cases of severe pancytopenia<br>○ Hepatobiliary system: amoebic liver abscess and spontaneous bacterial peritonitis<br>○ Gastrointestinal tract infections: enteric fever, dysentery, worm infestation, and abdominal tuberculosis.<br>○ Respiratory tract infections: lower respiratory tract infections, aspiration pneumonia, acute exacerbation of chronic obstructive pulmonary disease, atypical pneumonia, aspergillosis, and pulmonary tuberculosis<br>○ Urinary tract infections: lower urinary tract infections, catheter associated urinary tract infections, complicated & uncomplicated pyelonephritis, and candiduria<br>○ Central nervous system infections: meningitis & encephalitis (bacterial/viral/listeria/scrub/tubercular), and neurocysticercosis<br>○ Skin and soft tissue infections: cellulitis and infected diabetic foot<br>○ Other dermatological indications: tinea, scabies, hansen disease, oral candidiasis, and bedsores |  |  |  |  |  |  |

**Table 2:** Frequency of non-adherent categories of antimicrobial prescriptions.

|  |  |  |  |  |
| --- | --- | --- | --- | --- |
| **Non-adherent categories** | **Over prescription<sup>#</sup>** | **Under prescription<sup>\*</sup>** | **Choice not correct<sup>\$</sup>** | **Antimicrobial not indicated<sup>@</sup>** |
| Indicated surgical prophylaxis | 1 |  |  |  |
| Indicated medical prophylaxis | 2 | 2 |  | 6 |
| Bloodstream infections |  |  |  |  |
| Hepatobiliary system infections | 2 |  | 2 |  |
| Gastrointestinal system infections |  |  | 2 | 1 |
| Respiratory tract infections |  | 5 | 19 | 9 |
| Urinary tract infections | 4 | 3 | 11 | 2 |
| Central Nervous system infections |  | 1 |  | 2 |
| Skin & Soft tissue infections |  |  |  |  |
| Other Dermatological indications |  |  | 2 |  |
| Others† | 1 | 1 | 2 | 2 |
| Sepsis syndrome with defined focus |  |  | 8 | 8 |
| **Total** | **10** | **12** | **46** | **30** |
| **# Overprescription:** Those prescriptions where - either the administered dose of a given antimicrobial was higher than that recommended by the guidelines, and/ or total duration of antimicrobial therapy was greater than that recommended by the guidelines.<br>**\* Underprescription:** Those prescriptions where - either the administered dose of a given antimicrobial was lower than that recommended by the guidelines, and/ or total duration of antimicrobial therapy was shorter than that recommended by the guidelines.<br>**\$ Choice not correct:** Those prescriptions where- there was a failure in choosing the correct choice of antimicrobial agent for that particular condition, partially or fully; and/ or failure to review the antimicrobial treatment when microbiological culture data became available.<br>**@ Antimicrobial not indicated:** Those prescriptions where antimicrobials use was not warranted and/ or those prescriptions where patient had already completed the entire length of therapy during the course of hospital stay but as a part of de-escalating therapy, antimicrobials were prescribed that were not required. | | | | |

## Discussion

This prospective hospital data record based study reveals that the antimicrobial choice was appropriate and matched the guideline in majority of admitted patients. Adherence was observed to the right drug, dose, route of delivery, and duration/de-escelation, a basic principle of ASP. Full adherence to all the above mentioned parameters was observed in 2/3^rd^ prescriptions. 100% adherence was seen for treatment against blood stream and skin and soft-tissue infections, while <50% in medical prophylaxis.

Antimicrobial stewardship refers to interventions designed to promote the optimum use of antimicrobial agents, including right diagnosis, drug choice, dose, and duration of administration as well as de-escalation (5Ds) as soon as appropriate. Reaching to the right diagnosis constitutes the first D and is also the most difficult one. This requires in depth knowledge of the clinical manifestations of the various infections and the proper utilization of the microbiology lab services. Once the right diagnosis is made, next step is initiating the right drug (second D) as mentioned in the guidelines/ textbooks but while keeping in mind the local susceptibility patterns and most frequent isolates. This may vary from hospital to hospital or even from one ward to another. Third D is about the right dose and delivery route and one must keep in mind that the same drug may have different doses/delivery route for infections involving different infection sites (for e.g. vancomycin for *Clostridium difficile* vs meningitis). The fourth D is about the right duration and is very important to minimize the chances of emergence of drug resistant organisms and at the same time, to minimize the side effects. Fifth D, de-escalation requires that the initial antimicrobial therapy should be narrowed or even discontinued as soon as possible based on the clinical response, culture, and susceptibility results in order to avoid the emergence of drug resistant organisms.

To achieve this, practicing physician has to follow standard treatment guideline (STG). Along with implementation of guidelines, adherence assessment is essential. A single centre study conducted in Namibia demonstrated compliance to the Namibia STG to be 62% and in Kuwait, full adherence was achieved only in 30.4% prescriptions (11,12). This lower level of compliance could be due to lack of formal monitoring system as well as lack of outpatient ASP.

However, although suboptimal, our adherence was higher than that reported by previous studies, which is usually around 40 % (5). This may be due to the fact that we avoided sticking to any one particular guideline for the choice of antimicrobials, as was done in most of the previous studies. We have included a wide range of clinical conditions in our study and it would likewise be unfair to label a treatment as non-adherent to guidelines just because the guidelines used were different. We were flexible in our approach and asked the treating physician regarding their source of reference and accepted the treatment as adherent as long as the guideline being referred to was from a reputed source and confirmed by investigators. Various studies across the globe show that 20-50% of all antimicrobials prescribed are either unnecessary or inappropriate (13,14). In studies conducted in China and Bangladesh, 63% and 50 % respectively of the antimicrobial selected to treat proven bacterial infections were deemed inappropriate (15,16). Long treatment duration and sub-therapeutic or sub-optimal dosages have been correlated with increase in selective resistance (17).

In our study, the maximum non-adherence was found in the choice of antimicrobial being incorrect. This is a cause of serious concern as in these times of widespread internet access, all the guidelines are available at the touch of a button and indicates a lack of awareness. The present study confirms 10% antimicrobial use that was unnecessary. This lower than expected non-indicated antibiotic prescriptions is probably due to the fact that we have included only in-patients (where proper assessment and discussion is feasible before initiating any treatment) unlike other studies which have taken OPD prescriptions. It shows that physicians err on the side of over-prescription rather than under-prescription when in doubt or when complete evaluation is not feasible as in OPDs.

Our study has its set of limitations as well. It was a single centre study conducted in a single clinical department. Hence the number of cases in different infection syndromes that have been included is very skewed (for example some infection syndromes have only 2 cases whereas others have 67). Future studies that include a larger sample size may be more representative of the magnitude of problem. Secondly, we have not evaluated the prescriptions for the correctness of diagnosis. Since the study was conducted in the Internal Medicine department of a tertiary care institute, it was assumed that most diagnoses would be correct as per the available evidences. Also the resources available at hand did not allow for a complete re-evaluation of each patient for the correctness of diagnosis. Thirdly there was no follow up involved and hence final treatment outcomes were not assessed.

The appropriate use of antimicrobials is an essential part of patient safety and deserves careful oversight and guidance. Given the association between antimicrobial use and the selection of resistant pathogens, the frequency of inappropriate antimicrobial use is often used as a surrogate marker for the avoidable impact on AMR. The combination of effective ASP with a comprehensive infection control program has been shown to limit the emergence and transmission of antimicrobial resistant bacteria.

To improve adherence to antimicrobial guidelines, more efficient quality measures should be developed and implemented which include availability of the guidelines on the hospital information system, monitoring antimicrobial misuse through repetitive audits, and continuous education of the physicians to raise their awareness of proper prescription. Local institute protocol based on national/international guidelines and textbooks should be revised and periodically updated to cover all conditions treatable with antimicrobials.

## Conclusion and Recommendations

The study demonstrates sub-optimal compliance to the STG on infectious diseases with respect to choosing right drug, dose, delivery route, and duration/de-escelation, although having lower prevalence of over-prescription, under-prescription, incorrect, or unnecessary antimicrobial uses when compared to studies from other developing countries. There is thus, an urgent need to mandate prospective audit and feedback, to improve ASP and reduce AMR in long run.

## Data Availability

Data is available with PI.

## Acknowledgements

Ms Anjali Chauhan helped in drafting the data.

## Transparency declarations

None

## Ethical approval

The study was approved by the Institutional Ethics Committee, All India Institute of Medical Sciences, Rishikesh, India.

